# Determination of glomerular filtration rate, a spin off after contrast-enhanced computed tomography among critically ill patients – proof of concept

**DOI:** 10.1101/2023.09.12.23295373

**Authors:** Bertil Kågedal, Anders Helldén, Dženeta Nezirevic Dernroth, Anders Andersen, Andreas Ekman, Mats Haglund, Bharti Kataria, Frida Oskarsson, Lovisa Tobieson, Åse Östholm Balkhed, Håkan Hanberger

## Abstract

**Background:** Recently, Gong et al. (Gong et al. 2022) showed, in nine heathy subjects, that plasma clearance of high doses of iohexol given as contrast enhanced computed tomography (CT) could be used for determination of glomerular filtration rate (GFR). We utilized high doses of iohexol from angiographic or other contrast enhanced CT given to critical ill patients for calculation of GFR_iohexol_ and compared these data with standard low dose iohexol GFR determinations.

**Method:** Patients at intensive care units (ICUs) in Southeast Sweden intended for radiographic investigations that included injection of 45-120 ml of iohexol (Omnipaque) were included, and the concentration of iohexol in plasma was measured by HPLC. Iohexol clearance was calculated by the method of Bröchner-Mortensen. The following days was iohexol clearance determined using the standard low dose of 5 mL of iohexol. Sixteen patients admitted to ICUs were included in this pilot study.

**Results:** GFR after high dosing of iohexol at contrast enhanced CT could be measured for all sixteen critically ill patients. Patients with normal or increased renal function had neglectable iohexol concentrations the day following the CT scan. There was excellent correlation between GFR determination with high and standard low iohexol dosing among these 6 patients. Ten patients had decreased renal function and delayed elimination of iohexol, thus was not GFR measurement with low dose iohexol possible to analyse the day after CT scan with high dose iohexol.

**Conclusion:** This pilot study showed that GFR can be measured after high doses of iohexol at enhanced CT and compare well with the standard low dose of iohexol clearance determinations.

## Introduction

When dosing of renally excreted drugs, the patient’s glomerular filtration rate (GFR) must be considered. Current estimated GFR (eGFR) methods, which are based on plasma concentrations of creatinine and cystatin C, are too uncertain to be relied on in patients needing intensive care (Delanaye, Melsom, et al. 2016).

Critical ill patients are frequently investigated by iohexol contrast enhanced computed tomography (CT). Iohexol plasma clearance has for years been the reference method for determination of GFR, but the iohexol dose used is only 5-10% of that used for contrast enhanced CT.

The reference method, low dose iohexol clearance (GFR_iohexol_), is seldom used as support for drug dosing in patients admitted to ICUs. The method is considered complicated, takes time to implement, requires accurate information on amount of injected iohexol, and exact time schedule for blood sampling (Delanaye, Ebert, et al. 2016; Delanaye, Melsom, et al. 2016; Levey and Inker 2016). According to guidelines, the earliest an examination with standard low dose dosing of iohexol can be carried out is 3-4 days after contrast X-ray, which prevents early knowledge from a more accurate GFR than that from plasma creatinine or cystatin C. However, we hypothesized that clearance of the iohexol given at the radiology department can be used for more rapid information of GFR_iohexol_. A prerequisite is that the pharmacokinetics of iohexol is not dose-dependent. Interestingly, this has been shown in earlier studies (Bäck, Krutzen, and Nilsson-Ehle 1988; Hu et al. 2013).

Recently, Gong et al. (Gong et al. 2022) showed that ten times higher dosages of iohexol gave the same clearance rates as the standard dosage of 5 mL iohexol in nine heathy subjects. The aim of this study was to investigate if the administration of iohexol in contrast enhanced CT, on clinical indication, in critically ill patients could be used for determination of GFR by plasma iohexol clearance determination.

## Material and Methods

We conducted a study on ICU patients in collaboration with the Radiology departments at three hospitals in southeast of Sweden. The iohexol preparation used at these departments were Omnipaque 350 mg iodine/mL (GE Healthcare). The radiology departments provided the information on iohexol dose and time for administration to the ICU where four blood samples were taken 2-4 hours after the injection for analysis of plasma iohexol concentration.

Standard clearance investigations were performed the following days with low dose iohexol of 5 mL Omnipaque 300 mg iodine/mL.

We analysed the concentration of iohexol in plasma by HPLC and calculated iohexol clearance by the method of Bröchner-Mortensen (Bröchner-Mortensen 1972). Patients with remaining high iohexol concentration in the pre-study sample (zero sample) day 2 and later would impact on the calculation of the standard low dose iohexol clearance studies and were exclude in the present report. This study was approved by the Swedish Ethical Review Authority No 2020-04197. The patients gave informed consent to the study.

## Results

Iohexol clearance after CT in sixteen critically ill patients were median (range) 102 mL/min (28 – 152). The correlation between iohexol clearance determined after CT (high dosing day 1; 33975-90600 mg iohexol) and low dosing days 2 to day 8 (3232 mg iohexol) was excellent, y = 1.0255x – 0.8807mL/min, R^2^ = 0.8687 (N=6), where x = clearance with low dose iohexol and y = iohexol clearance after CT (Figure 1). Patients with normal or increased renal function had neglectable pre-injection iohexol concentrations (zero sample) the day after the CT scan, and further on the day after the 5 mL injections. For patients with low GFR was GFR measured the day after CT scan based on elimination of the high dose iohexol, but for these patients was repeated GFR measurement the following day with the standard method not feasible due to remaining high concentration of iohexol.

**Figure 1.**
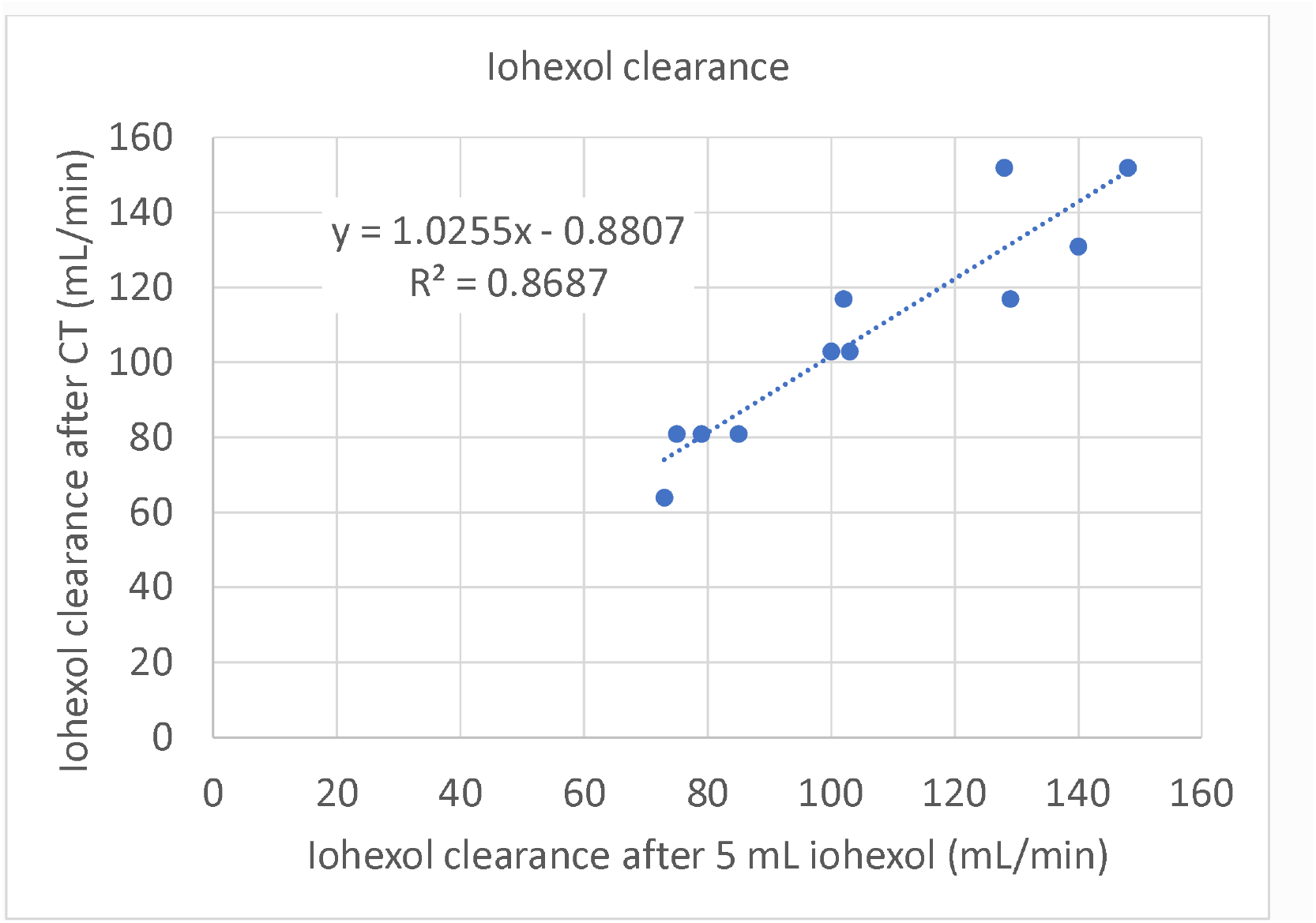
This figure shows the correlation between iohexol clearance measured with a low dose of 5 mL iohexol and iohexol clearance with high dose iohexol after computed tomography (CT) in six critically ill patients. Four patients had more than one low dose measurements. The figure contains data from patients with no concentrations of iohexol in the zero sample.

## Discussion

This pilot study shows that the results obtained after high dosing of iohexol at enhanced CT compare well with standard investigation with low dosing. Practically, the results obtained with the high iohexol dosing at radiology can be used for decision of dosing of drugs that are eliminated by glomerular filtration in ICU patients.

## Data Availability

All data produced in the present study are available upon reasonable request to the authors

## Notes

### Competing Interest Statement

The authors have declared no competing interest.

### Funding Statement

This study was funded by Medical Research Council of Southeast Sweden

### Author Declarations

Swedish Ethical Review Authority No 2020-04197

